# Clustering by Multiple Long-Term Conditions and Social Care Needs: A cohort study amongst 10,025 older adults in England

**DOI:** 10.1101/2023.05.18.23290064

**Authors:** Nusrat Khan, Christos V. Chalitsios, Yvonne Nartey, Glenn Simpson, Francesco Zaccardi, Miriam Santer, Paul Roderick, Beth Stuart, Andrew Farmer, Hajira Dambha-Miller

## Abstract

**Background:** People with Multiple Long-Term Conditions (MLTC) face health and social care challenges. This study aimed to classify people by MLTC and social care need (SCN) into distinct clusters and quantify the association between derived clusters and care outcomes.

**Methods:** A cohort study was conducted using the English Longitudinal Study of Ageing (ELSA), including people with up to ten MLTC. Self-reported SCN was assessed through 13 measures of difficulty with activities of daily living, ten measures of mobility difficulties, and whether health status was limiting earning capability. Latent class analysis was performed to identify clusters. Multivariable logistic regression quantified associations between derived SCN/MLTC clusters, all-cause mortality, and nursing home admission.

**Results:** The cohort included 9171 people at baseline with a mean age of 66·3 years; 44·5% were males. Nearly 70·8% had two or more MLTC, the most frequent being hypertension, arthritis, and cardiovascular disease. We identified five distinct clusters classified as high SCN/MLTC through to low SCN/MLTC clusters. The high SCN/MLTC included mainly women aged 70 to 79 years who were white and educated to the upper secondary level. This cluster was significantly associated with higher nursing home admission (OR = 8·97; 95% CI: 4·36 to 18·45). We found no association between clusters and all-cause mortality.

**Conclusions:** This results in five clusters with distinct characteristics that permit the identification of high-risk groups who are more likely to have worse care outcomes, including nursing home admission. This can inform targeted preventive action to where it is most needed amongst those with MLTC.

**What is already known on this topic:** While it is established that multiple long-term conditions are linked to an increased risk of hospitalisation, nursing home admission and mortality, no previous research has examined this risk in relation to clusters of MLTC and social care needs in England.

**What this study adds:** Using latent class analysis, this study identified five clusters by multiple long-term conditions and social care needs with distinct characteristics and quantified their relationship with nursing home admission and mortality.

**How this study might affect research, practice or policy:** The findings permit the identification of high-risk groups who are more likely to have worse care outcomes, including nursing home admission in the future. This can inform targeted preventive action to where it is most needed amongst those with MLTC. Recognition of MLTC and SCN clusters may also aid clinicians in moving away from a single disease management approach in older adults.

## Introduction

The growing burden of multiple long-term conditions (MLTC) is a significant global challenge for health and social care systems (1). MLTC is defined as the co-existence of two or more long-term conditions. One in four people worldwide is estimated to have MLTC, although prevalence rises with age, from 54% in those over 65 years of age to 83% in those over 85 years (2–4).

People living with MLTC require more intensive and complex person-centred care over a longer period than those with a single condition, which increases service utilisation and care costs due to the holistic nature of multiple diseases for specialised treatment requirements (5). Earlier studies have shown that those with MLTC aged between 50 to 64 years report difficulty with activities of daily living (ADL) and mobility in 15% and 18%, respectively (6). A recent analysis of the Health and Retirement Study in China found that nearly one-quarter of participants with MLTC developed difficulty with one or more ADL during middle age (7). MLTC also increases the likelihood of frailty, reduced mobility, and a general functional decline that often significantly impairs personal independence. In turn, this has increased the demand for social care, including higher levels of admissions to nursing or care homes, increased need for assisted living and a growth in ‘homecare’ support services to enable people to live independently as long as possible (8,9). Earlier studies have linked MLTC to an increased risk of hospitalisation, nursing home admission and mortality (10–13).

Given the growing numbers of people presenting with complex health and social care needs (SCN) and the increased burden of MLTC, clustering approaches could present a strategy for identifying those with specific combinations of MLTC and SCN who are at risk of increasing ill health or loss of independence, nursing home admission and/or death. Clustering relies on the fact that common conditions group together in predictable patterns within a population (14,15). Latent class analysis (LCA) has been a commonly used algorithm to identify clusters in cohorts of people with MLTC (16,17). Clustering by both MLTC and SCN may allow more precise identification of those who could benefit most from preventive interventions and increased resource allocation in a holistic way (10,18). Although some advances have been made in MLTC clustering research (19,20), there is a scarcity of evidence considering SCN in combination with MLTC (21,22). This study aimed to classify people by MLTC and SCN into distinct clusters and quantify the association between derived clusters and care outcomes.

## Methods

### Data source

The English Longitudinal Study of Aging (ELSA) is a cohort study of people aged 50 years or older living in England (23). Details of the study have been reported elsewhere but in brief, a population-representative sample of members was drawn from the Health Survey for England (HSE) from 2002, with repeated waves of follow-up every two years and additional nurse visits to assess biomarkers every four years (23,24). It included 12099 people in 2002 as the study entry point, with a wide range of data collected on physical and mental health, well-being, finances, and attitudes around ageing over time. ELSA is an open cohort, and refreshment samples have been added by corresponding HSE surveys depending on the proportional age requirement for ELSA (e.g., 50 to 74 years and their partners for wave 4 and 50 to 53 years and partners for wave 9), using cross-sectional and longitudinal weights for the core surveyed. The datasets of ELSA harmonised (elsa_harmonised) and ELSA harmonised G2 (elsa_harmonised_g2) were used for this study.

### Study design and population

This cohort study uses ELSA wave 2 (2002/3) to wave 9 (2018/19), with or without MLTC. Our baseline was wave□2, which included data from nurse visits, allowing more MLTC to be included and verified by nurse records rather than relying on self-reported data. This study was conducted, and findings were reported in line with the STROBE guidelines for observational studies using routinely collected health data (checklist reported in the **Supplementary Material**) (24).

### Multiple Long-Term Conditions

Ten MLTC were available in this dataset based on our previous works and consensus on defining MLTC, which identified a total of 59 disease conditions (25–27). The ten conditions available in ELSA included hypertension, diabetes, cancer, lung disease, cardiovascular disease, stroke, mental health disorders, arthritis, Parkinson’s disease and dementia. The presence of these conditions is defined in ELSA by self-reporting the last two years and nurse review of healthcare records (23). Due to the small sample size (less than ten cases), some conditions were combined following clinical discussion and consensus: depression was included among mental health disorders, asthma within lung disease, Alzheimer’s disease within dementia, heart attack, angina, heart murmur, abnormal heart rhythm, and congestive heart failure within cardiovascular diseases. We considered the highest number of MLTCs developed by each participant across multiple waves.

### Social Care Needs

SCN variables were identified by a parallel Delphi consensus study that included professionals, people living with MLTC, and informal carers identifying SCN in MLTC (26,27). Variables identified from the Delphi were mapped to the ELSA data dictionary, resulting in an operational definition of SCN as follows: (i) 13 self-report (yes/no) difficulties in ADL; (ii) ten self-report binary (yes/no) difficulty in physical mobility; (iii) self-report on whether an individual’s health status was limiting earning capability (28). The ELSA questionnaire included standardised measures for quantifying ADL and mobility variables, which have undergone extensive validation in previous studies (29). The ADL variables included: difficulty with dressing; putting on shoes and socks; walking across a room; bathing or showering; eating such as cutting up food; getting in and out of bed; using the toilet; getting up or down; using a map for location; preparing a hot meal; shopping for groceries; making telephone calls;taking medications; doing work around house and garden; managing money, e.g. paying bills, and keeping track of expenses. The mobility variables included: difficulty in the ability to walk 100 yards; sit for 2 hours; get up from the chair after sitting for prolonged periods; climb several flights of stairs without resting; climb one flight of stairs without resting; kneel or crouch; reach or extend arms above shoulder level; pull or push large objects; lift or carry weights over 10 pounds; picking up 5p coin from a table. Health status limiting earning capability was a variable that was also included under our definition of SCN. It denotes whether an impairment or health problem limits the type or amount of paid employment (23,28). We combined the 13 items of ADL and the ten items of difficulty with physical mobility into one composite score for ADL and mobility. For each item, a score was assigned for the absence and presence of ADL and mobility difficulties, respectively (score 0 if absence and score 1 if presence).Therefore, the overall sum of scores across all items was either 0 or ≥1. Those with a sum of ≥1 were considered to have ADL or mobility difficulties. For our SCN variable, we considered the maximum number of SCN developed by each participant during the study period.

### Care outcomes

The outcomes of interest were nursing home admission and all-cause mortality in the previous two years. These were self-reported with end-of-life or after-death interviews on waves 2, 3, 4 and 6 among a sample of family members or carers of ELSA participants who had recently passed away, asking about the circumstances around the respondent’s final stages of life (23,24).

### Socio-demographic

Self-reported information was available at baseline for age (continuous), sex, and ethnicity (grouped within the database as whites or non-whites). Age was further categorised for analysis (50-59, 60-69, 70-79, ≥80 years old). Education level was categorised into four groups: less than upper secondary level, upper secondary and vocational level, tertiary level, and others. Employment status was categorised as working for payment and not working for payment. Marital status was categorised into three groups: never married, married/having a partner and separated/divorced/widowed. To minimise the impact of missing data, we used data provided in the subsequent waves for any missing information at baseline.

### Statistical analysis

We summarised the characteristics of the cohort using descriptive statistics comparing individuals with and without MLTC. LCA was conducted to identify distinct clusters of MLTC and SCN. LCA is a model-based clustering technique that classifies individuals into clusters based on multiple characteristics in a cohort (in this case, MLTC and SCN). The posterior probability of belonging to each cluster can be obtained for each participant; assigned according to their highest probability of membership. The underlying assumption of LCA is that individuals belong to unobserved (latent) clusters but can be classified based on information available in observed data through a likelihood function. A series of latent class models were fitted iteratively, beginning with 2 clusters and up to 6 clusters. Six clusters were the maximum fitted to balance optimal fit with clinical utility. The optimal number of latent clusters was determined using the dissimilarity index and the Bayesian Information Criterion (BIC) as robust indicators of the cluster alongside clinical interpretation (30,31). BIC was used to compare several plausible models with the lowest values to indicate the best-fitting model. Multivariable logistic regression was computed to assess the association of each MLTC/SCN cluster with the outcomes (nursing home admission and all-cause mortality), adjusted for age, sex, ethnicity, marital status, education, and employment.The cluster with the highest number of people was considered as the reference category. Data management and analyses were conducted using Stata M.P. (version 17).

## Results

### Descriptive characteristics

A total of 9171 people were identified at baseline (wave 2). They were mainly white (98%) and female (55·5%), with a mean (SD) age of 66·3 (10) years (**Table 1**). Amongst them, the MLTC cohort comprised 70.8% (**Table 1**). Most were married or partnered (66·4%), 11.2% completed level 3 upper education, and nearly two-thirds (72·9%) were not working. At baseline, 36·8% of those with MLTC had at least one ADL difficulty, and 68·6% had at least one mobility difficulty (**Table 1**).

**Table 1.** Characteristics at baseline (wave 2).

|  | **Total (N=9171)** | **MLTC**<br>**(6489, 70.8%)** | **No MLTC**<br>**(2682, 29.2%)** |
| --- | --- | --- | --- |
| **Age, years, mean (SD)** | 66.3 (10) | 67.5 (9.8) | 63.4 (9.7) |
| **Age, years** |  |  |  |
| 50-59 | 2925 (31.9) | 1739 (26.8) | 1186 (44.2) |
| 60-69 | 2920 (31.8) | 2091 (32.2) | 829 (30.9) |
| 70-79 | 2203 (24) | 1775 (27.3) | 428 (16) |
| ≥80 | 1123 (12.2) | 884 (13.6) | 239 (8.9) |
| **Sex** |  |  |  |
| Male | 4084 (44.5) | 2791 (43) | 1293 (48.2) |
| Female | 5087 (55.5) | 3698 (57) | 1389 (51.8) |
| **Ethnicity** |  |  |  |
| White | 8963 (98) | 6341 (97.7) | 2622 (97.8) |
| Non-white | 206 (2) | 148 (2.3) | 58 (2.2) |
| Missing* | 2 (0.02) | 0 (0) | 2 (0.1) |
| **Marital status** |  |  |  |
| Married/Partnered | 6335 (69.1) | 4308 (66.4) | 2027 (75.6) |
| Separated/Divorced/Widowed | 2411 (26.3) | 1890 (29.1) | 521 (19.4) |
| Never married | 424 (4.6) | 291 (4.5) | 134 (5) |
| Missing* | 1 (0.01) | 0 (0) | 1 (0.04) |
| **Education\*\*** |  |  |  |
| Less than upper secondary | 3563 (38.8) | 2681 (41.3) | 882 (32.9) |
| Upper secondary and vocational training | 3687 (40.2) | 2483 (38.3) | 1204 (44.9) |
| Tertiary education | 1124 (12.3) | 724 (11.2) | 400 (14.9) |
| Other | 784 (8.5) | 596 (9.2) | 188 (7) |
| Missing* | 13 (0.1) | 5 (0.1) | 8 (0.3) |
| **Employment** |  |  |  |
| Currently working | 3075 (33.5) | 1755 (27.1) | 1320 (49.2) |
| Not working | 6095 (66.5) | 4734 (72.9) | 1361 (50.7) |
| Missing* | 1 (0.01) | 0 (0) | 1 (0.04) |
| **Long-term conditions** |  |  |  |
| Diabetes | 744 (8.1) | 718(11.1) | 26(1.0) |
| Hypertension | 3793 (41.3) | 3396(52.3) | 397(14.8) |
| Cancer | 662 (7.2) | 607(9.4) | 62(2.3) |
| Lung diseases | 1503 (16.4) | 1387(21.4) | 116(4.3) |
| Cardiovascular diseases | 1981 (21.6) | 1851(28.5) | 130(4.9) |
| Stroke | 442 (4.8) | 429(6.6) | 13(0.5) |
| Mental health disorders | 1964 (21.4) | 1818(28.0) | 146(5.5) |
| Arthritis | 3207 (35) | 2919(45.0) | 288(10.7) |
| Parkinson disease | 54 (0.6) | 50(0.8) | 4(0.2) |
| Dementia | 77 (0.7) | 70(1.1) | 7(0.3) |
| **Social care needs** |  |  |  |
| Difficulties in any ADL | 2716 (29.6) | 2390 (36.8) | 326(12.2) |
| Difficulty in any physical mobility | 5423 (59.1) | 4454 (68.6) | 969(36.2) |
| Health status limiting earning capability | 2990 (32.6) | 2620 (40.4) | 370(14.0) |

All figures are expressed as absolute numbers and percentages unless otherwise specified.
\*missing = missing + not available + no respondent
\*\*Upper secondary = level 3 secondary education. Typically aged 16-18 years

A total of 654 individuals died during the follow-up period (all-cause mortality 3·9%), with 24·9% (N=163) having stayed in a nursing home. Complete data were available for 10025 (59·9%) people with MLTC from wave 2 to wave 9.

### Clustering MLTC and SCN

We applied LCA in a total of 10025 participants and, based on the lowest BIC (**Supplementary Table 1**), identified five distinct clusters (**Figure 1 & Supplementary Table 2**). The dissimilarity index was 0·25. Cluster 1 (9·3%, N = 934) represented the highest probability of hypertension (81%), cardiovascular disease (34%), and mental health disorder (37%). In cluster 2 (13·7%, N = 1370), 85% of people had a high probability of mobility difficulty, followed by arthritis, mental health disorders, and cardiovascular diseases. Cluster 3 (21·9%, N = 2197) was dominated by a high probability of SCN conditions, with 98% of mobility difficulties and 49% with health status limiting work. Cluster 4 (49·2%, N = 4937) was also dominated by ADL difficulties with a probability of 98%, followed by 75% of arthritis and 67% of hypertension. However, cluster 5 (5·9%, N = 587) was prominently dominated by all the SCN, with a 99% probability of ADL difficulties, 98% of mobility difficulties, and 80% of health status limiting earning capability. All the clusters were dominated by arthritis, mental health disorders, cardiovascular diseases, and hypertension in terms of disease conditions.

**Figure 1.**
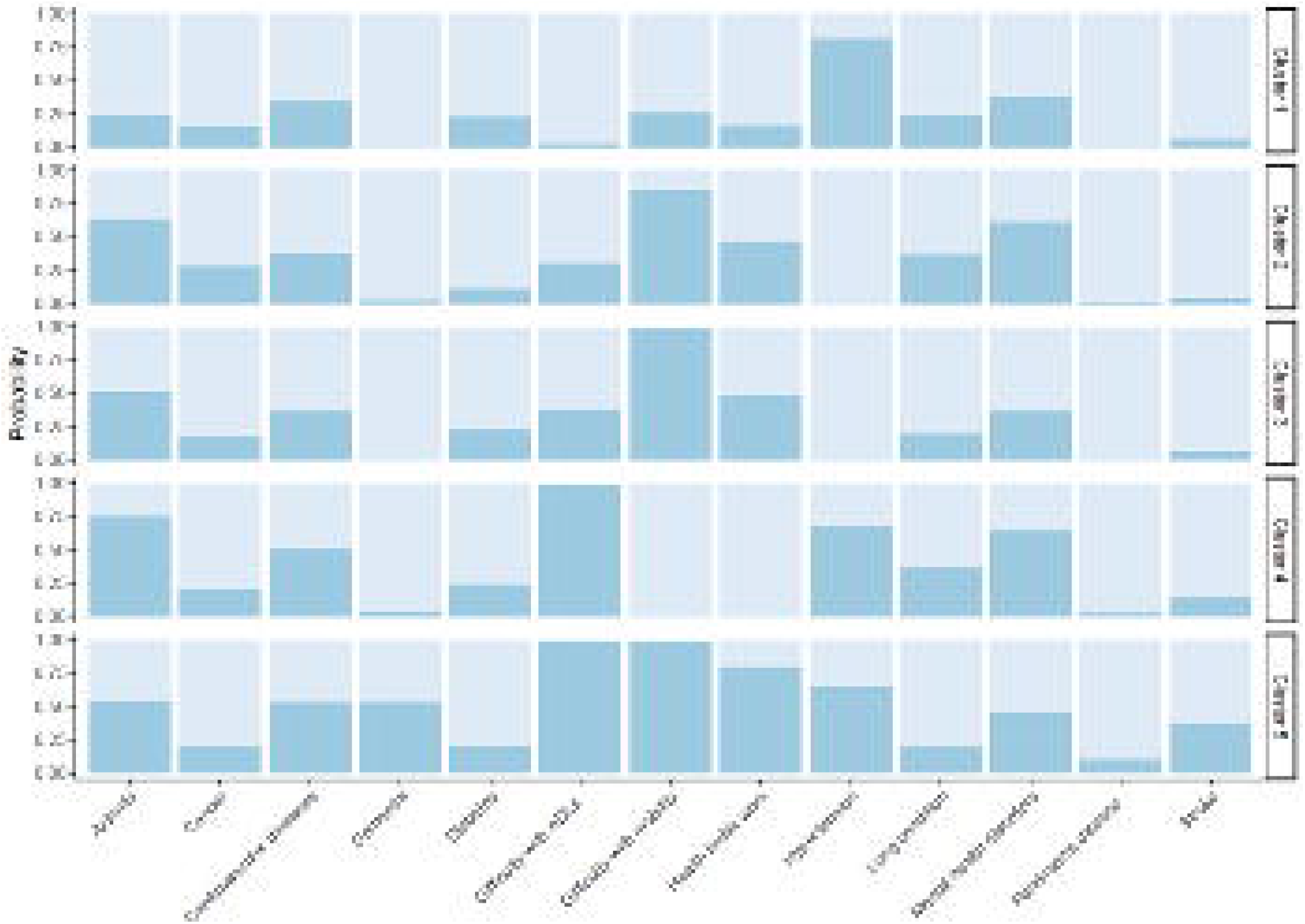
Item response probabilities of multiple long-term conditions and social care needs clusters.

In terms of socio-demographic characteristics of the clusters; cluster 1 had the youngest median (IQR) age of 57 (53 to 65) years (**Table 2**). All the clusters had more females than males except for cluster 1 (64% vs 36%). Individuals in cluster 5 (dominated by all three SCN variables) had the highest median age (IQR) of 75 years (69 to 81). The clusters differed by marital status and education; specifically, from low SCN/MLTC cluster 1 to high SCN/MLTC cluster 5, the proportion of separated/divorced/widowed people increased from 13·5% to 32·5%. A trend of a lower educational level was observed with progression from low SCN to high SCN. Only 23% of individuals in the low SCN/MLTC cluster 1 had less than upper secondary level education compared to nearly 50% in the high MLTC/SCN cluster 5. Another low SCN/MLTC cluster (cluster 4) had a very high proportion of individuals who were previously married (31%) and unemployed (79%). About 21·5% of the individuals received tertiary education in cluster 1 and 18·5% in cluster 2 compared to a much lower 8·7% in cluster 5.

**Table 2:** Socio-demographic characteristics by multiple long-term conditions and social care needs clusters

| Characteristics | Cluster 1 (n=934;<br>9.3%) | Cluster 2 (n=1370;<br>13.7%) | Cluster 3 (2197;<br>21.9%) | Cluster 4 (4937;<br>49.2%) | Cluster 5 (n=587;<br>5.9%) |
| --- | --- | --- | --- | --- | --- |
| **Age(years), median (IQR)** | 57 (53 – 65) | 59 (54 - 67) | 61 (56 - 69) | 66 (58 - 74) | 75 (69 - 82) |
| **Age (years)** |  |  |  |  |  |
| 50-59 | 182 (41.2) | 288(37.7) | 479(34.4) | 763(22.2) | 27(5.9) |
| 60-69 | 166(37.6) | 285(37.4) | 486(34.9) | 1065(31.0) | 89(19.3) |
| 70-79 | 83(18.8) | 143(18.7) | 312(22.4) | 1056(30.8) | 181(39.3) |
| 80+ | 11(2.5) | 47(6.2) | 117(8.4) | 546(15.9) | 163(35.4) |
| **Sex** |  |  |  |  |  |
| Male | 596(63.7) | 570(41.6) | 1021(46.5) | 2086(42.3) | 262(44.6) |
| Female | 339(36.3) | 800(58.4) | 1176(53.5) | 2851(57.7) | 325(55.4) |
| **Ethnicity** |  |  |  |  |  |
| White | 879 (94.0) | 1320(96.4) | 2104(95.8) | 4759(96.4) | 573(97.6) |
| Non-white | 56(6.0) | 50(3.6) | 93(4.2) | 177(3.6) | 14(2.4) |
| **Marital status** |  |  |  |  |  |
| Married/ Partnered | 763 (81.6) | 1034(75.5) | 1638(74.6) | 3106(62.9) | 374(63.7) |
| Separated/ Divorced/ Widowed | 126(13.5) | 262(19.1) | 449(20.4) | 1537(31.1) | 194(33.0) |
| Never married | 46(4.9) | 73(5.3) | 110(5.0) | 294(6.0) | 19(3.2) |
| **Education** |  |  |  |  |  |
| Less than upper secondary | 213(23.3) | 345(25.4) | 687(31.6) | 2221(45.4) | 284(48.8) |
| Upper secondary and vocational training | 453 (49.4) | 657(48.3) | 961(44.2) | 1842(37.6) | 195 (33.5) |
| Tertiary education | 197(21.5) | 252(18.5) | 352(16.2) | 407(8.3) | 54(9.3) |
| Other | 54(5.9) | 106(7.8) | 172(7.9) | 427(8.7) | 49(8.4) |
| **Employment** |  |  |  |  |  |
| Not working | 354(37.9) | 612(44.7) | 1159(52.8) | 3891(78.8) | 534(91.0) |
| Currently working | 580(62.1) | 757(55.3) | 1038(47.2) | 1045(21.2) | 53(9.0) |

The numbers are presented as absolute numbers and percentages unless otherwise specified.

### MLTC/SCN clusters and care outcomes

Cluster 5 had higher odds of nursing home admission (aOR = 8·97; 95%CI 4·36 to 18·45), and none of the clusters was associated with all-cause mortality compared to cluster 4 (**Figure 2**).

**Figure 2:**
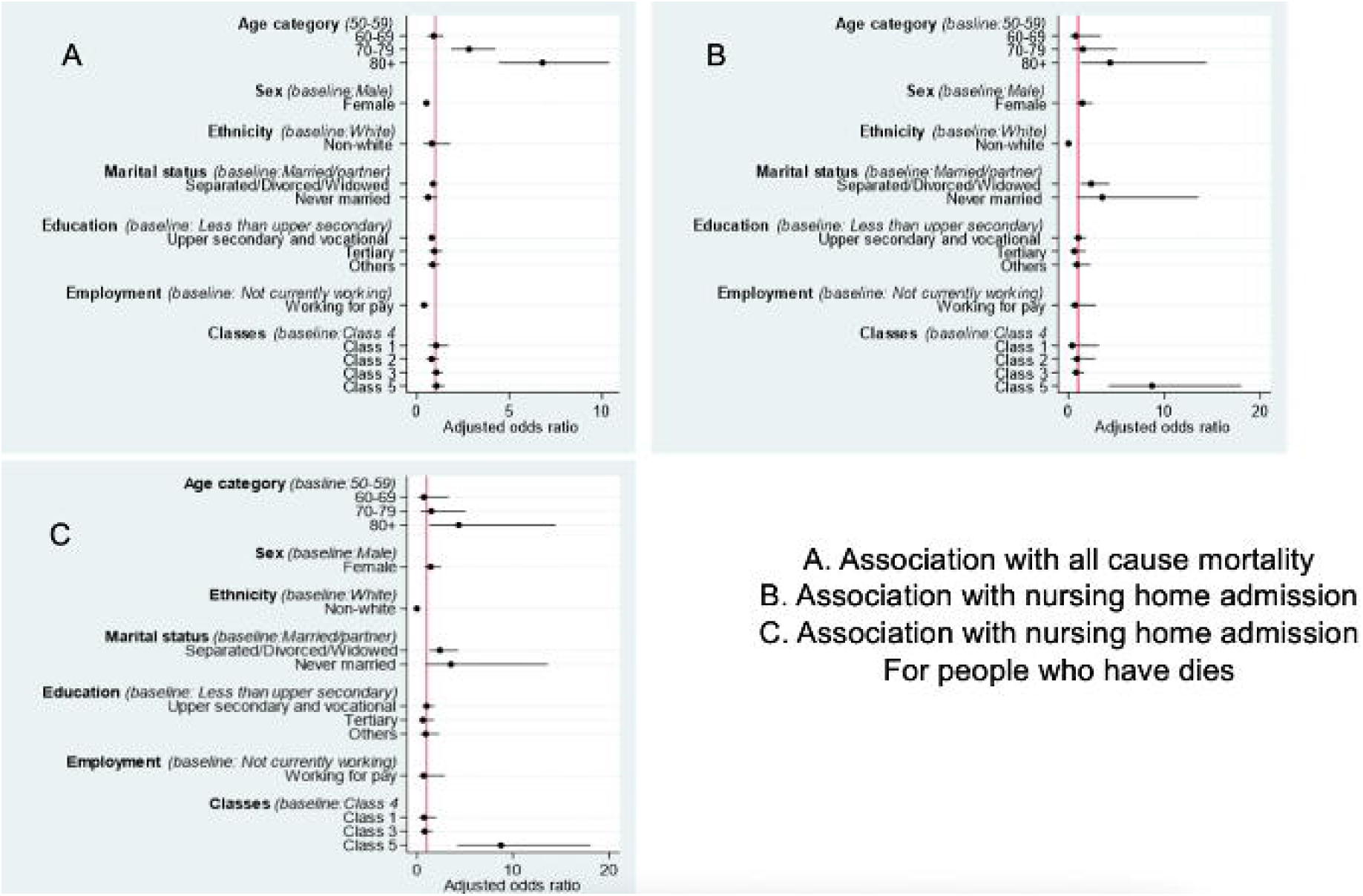
Odds ratios (with 95% CI) of the association between clusters of multiple long-term conditions and social care needs and the outcomes of interest. The estimates are adjusted for all covariates shown in the forest plot.

## Discussion

We established five distinct MLTC/SCN clusters amongst 10,025 older adults in England. Amongst the ten conditions that were examined, only a combination of arthritis, mental health disorders, cardiovascular diseases, and hypertension dominated when combined with SCN. This specific combination of MLTC and high SCN (cluster 5) was associated with a higher risk of nursing home admission than the reference category after adjusting for confounders, including age. People with a lower level of education, unemployed, or separated/divorced/widowed were more likely to fall into this cluster.

One of the major current and upcoming difficulties for healthcare systems globally has been identified as MLTC (32). To tailor health care design, broad general descriptions of the health outcomes and demands of patients with MLTC (i.e., based on counts of conditions) are not helpful. As a result, there have recently been calls to shift away from merely counting diseases in favour of a more specialised comprehension of which medical problems are most likely to co-occur (e.g., clustering of diseases). Therefore, to deliver optimal care to possible homogenous patient population groups, there has been increasing interest in focusing research on clinically meaningful clusters and wider determinants instead of considering MLTC as a general concept. (32–34).

Clustering by social care or wider health determinants remains limited in the present literature (35). Most observational research to date in MLTC has focused on the co-occurrence of conditions and biological determinants (30,31,36,37). Two previous systematic reviews summarise the literature on MLTC clusters and highlight that this is primarily described by the co-occurrence of conditions, including cardiovascular diseases and mental health disorders (36,38). We also observed this in our findings, although this was additionally accompanied by arthritis and hypertension. A recent longitudinal study of 16 years among older adults in Taiwan revealed that the cardiometabolic MLTC pattern had a much stronger association with increased mortality (39). A recent comparative study on MLTC clusters in the USA, Canada, England and Ireland showed the patterns of disease clusters and the risk factors related to each disease cluster were similar; however, the probabilities of the diseases within each cluster differed across countries (39). This highlights the necessity of identifying different clusters of MLTC and conditions with high probabilities to cooccurrence (39,40). MLTC in general, has been explored widely to identify sociodemographic risk factors. Distinct sociodemographic characteristics of MLTC clusters have also been identified in a limited number of studies. In a large-scale 16-year longitudinal study in Brazil, women and men presented different mortality patterns according to MLTC combinations (41). Four longitudinal studies from electronic health records in the US, UK, Europe, and China highlighted the role of marital relationships in shaping the trajectory of health and well-being across the life course in people with MLTC (42). However, the generalisation of MLTC research findings in various contexts is complex, given the multimodal nature.

To our knowledge, this is the first study that has examined clustering by both MLTC and SCN in England. Our study has a number of strengths.ELSA is an established longitudinal cohort with limited missing values and a relatively high follow-up. ELSA is also nationally representative of people aged 50 years and older. We also included multiple measures of SCN for a more reliable understanding of this concept. Our statistical analysis was accompanied by multiple layers of clinical discussion and interpretation in deriving meaningful clusters. Some limitations of our study should also be acknowledged. Firstly, we used cross-sectional data; therefore, causality cannot be inferred. Secondly, many of the variables were derived through self-report health and social care assessment, which may be subject to information and recall bias. The analysis used only ten MLTC based on what was available in the ELSA data, so a different association might have arisen if other MLTC or SCN had been considered. Additionally, when interpreting the results of observational studies, the sample might only represent healthy survivors in the population. Finally, the reverse causality between MLTC and ADL and mobility difficulties could not be addressed, although there is an abundance of previous literature reporting on this direction of association (8,29,43).

Current literature calls for more work on holistic clusters considering wider determinants to deliver optimal care. Our clustering approach and findings may offer a unique solution highlighting those with specific combinations of MLTC and SCN who are at risk of worse care outcomes, including nursing home admission. This has important policy implications as it may allow more precise identification of those who could benefit most from preventive measures (16,35,36). Previous studies have shown that older people with MLTC and social needs are likely to have worse health outcomes (31,34), but our data provide more specific combinations of conditions which were statistically and socio-demographically distinct. Identifying target populations with complex MLTC clusters can further build better health and social care system models and interventions that better integrate the clinical management of MLTC while concurrently addressing SCN.

In conclusion, this study demonstrated the feasibility of classifying people by MLTC and SCN clusters and shows that this approach could be clinically meaningful. We identified SCN/MLTC clusters with varying health and social demand and were able to differentiate between clusters by socio-demographic characteristics. We also showed that care outcomes could vary by cluster. Further research will need to explore the temporality of these associations and examine long-term outcomes beyond nursing home admission and mortality, including economic analysis.

## Supporting information

Strobe Statement

## Data Availability

ELSA data were available through the UK Data Archive and are widely available to access in this way; as such, our study data will not be made available for access.

## Declarations

### Ethical Approval

Ethical approval was granted by the University of Southampton Faculty of Medicine Research Committee (67953).

### Consent to participate

Secondary data has been used in this analysis as available from the ELSA database upon institutional request and approvals.

### Consent to publish

The authors affirm using secondary data for the analysis with no direct involvement of human research participants. As per the ELSA code and guidelines, this is a publication with anonymous participant information.

### Author Contributions

All authors contributed to the study’s conception and design. Data management and analysis were performed by HDM, YN and NK. The draft of the manuscript was written by NK, CVC and all authors commented on subsequent versions of the manuscript. All authors read and approved the final manuscript.

## Acknowledgement

We would like to thank Ms Firoza Davies for her contribution as a patient and public representative in this study.

## References

1. Multimorbidity: a priority for global health research. 2018;

2. Kingston A, Robinson L, Booth H, Knapp M, Jagger C, Adelaja B, et al. Projections of multi-morbidity in the older population in England to 2035: Estimates from the Population Ageing and Care Simulation (PACSim) model. Age Ageing. 2018 May 1;47(3):374–80.

3. Van Zon SKR, Reijneveld SA, Galaurchi A, Mendes De Leon CF, Almansa J, Bültmann U. Multimorbidity and the Transition Out of Full-Time Paid Employment: A Longitudinal Analysis of the Health and Retirement Study. Journals Gerontol Ser B [Internet]. 2020 Feb 14 [cited 2022 Dec 11];75(3):705–15. Available from: https://academic.oup.com/psychsocgerontology/article/75/3/705/5488785

4. Nunes BP, Flores TR, Mielke GI, Thumé E, Facchini LA. Multimorbidity and mortality in older adults: A systematic review and meta-analysis. Arch Gerontol Geriatr [Internet]. 2016 Nov 1 [cited 2022 Dec 11];67:130–8. Available from: https://pubmed.ncbi.nlm.nih.gov/27500661/

5. Coste J, Valderas JM, Carcaillon-Bentata L. Estimating and characterizing the burden of multimorbidity in the community: A comprehensive multistep analysis of two large nationwide representative surveys in France. PLOS Med [Internet]. 2021 Apr 1 [cited 2022 Dec 11];18(4):e1003584. Available from: https://journals.plos.org/plosmedicine/article?id=10.1371/journal.pmed.1003584

6. Gardener EA, Huppert FA, Guralnik JM, Melzer D. Middle-aged and mobility-limited prevalence of disability and symptom attributions in a national survey. J Gen Intern Med 2006 2110 [Internet]. 2006 Oct [cited 2022 Dec 11];21(10):1091–6. Available from: https://link.springer.com/article/10.1111/j.1525-1497.2006.00564.x

7. Wang Z, Peng W, Li M, Li X, Yang T, Li C, et al. Association between multimorbidity patterns and disability among older people covered by long-term care insurance in Shanghai, China. BMC Public Health [Internet]. 2021 Dec 1 [cited 2022 Dec 11];21(1):1–10. Available from: https://bmcpublichealth.biomedcentral.com/articles/10.1186/s12889-021-10463-y

8. Green I, Stow D, Matthews FE, Hanratty B. Changes over time in the health and functioning of older people moving into care homes: analysis of data from the English Longitudinal Study of Ageing. Age Ageing [Internet]. 2017 Jul 1 [cited 2023 Feb 4];46(4):693–6. Available from: https://academic.oup.com/ageing/article/46/4/693/3572451

9. Head A, Fleming K, Kypridemos C, Pearson-Stuttard J, O’Flaherty M. Multimorbidity: the case for prevention. J Epidemiol Community Health [Internet]. 2021 Mar 1 [cited 2023 Jan 20];75(3):242–4. Available from: https://pubmed.ncbi.nlm.nih.gov/33020144/

10. Nunes BP, Flores TR, Mielke GI, Thumé E, Facchini LA. Multimorbidity and mortality in older adults: A systematic review and meta-analysis. Arch Gerontol Geriatr [Internet]. 2016 Nov 1 [cited 2023 Feb 4];67:130–8. Available from: https://pubmed.ncbi.nlm.nih.gov/27500661/

11. Cassell A, Edwards D, Harshfield A, Rhodes K, Brimicombe J, Payne R, et al. The epidemiology of multimorbidity in primary care: a retrospective cohort study. Br J Gen Pract [Internet]. 2018 Apr 1 [cited 2022 Dec 11];68(669):e245. Available from: /pmc/articles/PMC5863678/

12. Doessing A, Burau V. Care Coordination of Multimorbidity: A Scoping Study. J Comorbidity. 2015 Jan;5(1):15–28.

13. Price ML, Surr CA, Gough B, Ashley L. Experiences and support needs of informal caregivers of people with multimorbidity: a scoping literature review. https://doi.org/101080/0887044620191626125 [Internet]. 2019 Jan 2 [cited 2022 Dec 11];35(1):36–69. Available from: https://www.tandfonline.com/doi/abs/10.1080/08870446.2019.1626125

14. Whitson HE, Johnson KS, Sloane R, Cigolle CT, Pieper CF, Landerman L, et al. Identifying Patterns of Multimorbidity in Older Americans: Application of Latent Class Analysis. J Am Geriatr Soc [Internet]. 2016 Aug 1 [cited 2022 Dec 11];64(8):1668–73. Available from: https://onlinelibrary.wiley.com/doi/full/10.1111/jgs.14201

15. Smith SM, Wallace E, O’Dowd T, Fortin M. Interventions for improving outcomes in patients with multimorbidity in primary care and community settings. Cochrane Database Syst Rev. 2021 Jan 15;2021(1).

16. Park B, Lee HA, Park H. Use of latent class analysis to identify multimorbidity patterns and associated factors in Korean adults aged 50 years and older. PLoS One [Internet]. 2019 Nov 1 [cited 2023 Jan 20];14(11):e0216259. Available from: https://journals.plos.org/plosone/article?id=10.1371/journal.pone.0216259

17. Larsen FB, Pedersen MH, Friis K, Gluèmer C, Lasgaard M. A Latent Class Analysis of Multimorbidity and the Relationship to Socio-Demographic Factors and Health-Related Quality of Life. A National Population-Based Study of 162,283 Danish Adults. PLoS One [Internet]. 2017 Jan 1 [cited 2023 Jan 20];12(1):e0169426. Available from: https://journals.plos.org/plosone/article?id=10.1371/journal.pone.0169426

18. Nuño-Solínis R, Ponce S, Urtaran-Laresgoiti M, Lázaro E, Errea Rodríguez M. Factors Influencing Healthcare Experience of Patients with Self-Declared Diabetes: A Cross-Sectional Population-Based Study in the Basque Country. Healthc (Basel, Switzerland) [Internet]. 2021 Apr 28 [cited 2022 Dec 11];9(5). Available from: http://www.ncbi.nlm.nih.gov/pubmed/33925113

19. Gaulin M, Simard M, Candas B, Lesage A, Sirois C. Combined impacts of multimorbidity and mental disorders on frequent emergency department visits: A retrospective cohort study in Quebec, Canada. CMAJ. 2019;191(26):E724–32.

20. Whitty CJM, Watt FM. Map clusters of diseases to tackle multimorbidity. Nature. 2020 Mar 26;579(7800):494–6.

21. Simpson G, Stuart B, Hijryana M, Akyea RK, Stokes J, Gibson J, et al. Eliciting and prioritising determinants of improved care in Multiple Long Term Health Conditions (MLTC): A modified online Delphi study. medRxiv [Internet]. 2023 Mar 19 [cited 2023 Mar 27];2023.03.19.23287406. Available from: https://www.medrxiv.org/content/10.1101/2023.03.19.23287406v1

22. Dambha-Miller H, Simpson G, Akyea RK, Hounkpatin H, Morrison L, Gibson J, et al. Development and Validation of Population Clusters for Integrating Health and Social Care: Protocol for a Mixed Methods Study in Multiple Long-Term Conditions (Cluster-Artificial Intelligence for Multiple Long-Term Conditions). JMIR Res Protoc [Internet]. 2022 Jun 1 [cited 2023 Feb 19];11(6). Available from: https://pubmed.ncbi.nlm.nih.gov/35708751/

23. Steptoe A, Breeze E, Banks J, Nazroo J. Cohort profile: the English longitudinal study of ageing. Int J Epidemiol [Internet]. 2013 Dec [cited 2022 Dec 11];42(6):1640–8. Available from: https://pubmed.ncbi.nlm.nih.gov/23143611/

24. Cadar D, Abell J, Matthews FE, Brayne C, David Batty G, Llewellyn DJ, et al. Cohort Profile Update: The Harmonised Cognitive Assessment Protocol Sub-study of the English Longitudinal Study of Ageing (ELSA-HCAP). Int J Epidemiol [Internet]. 2021 Jun 1 [cited 2022 Dec 11];50(3):725–726I. Available from: https://pubmed.ncbi.nlm.nih.gov/33370436/

25. Simpson G, Stokes J, Farmer A D-MH. Social care needs in multimorbidity. J R Soc Med. 2023;

26. Simpson G, Stuart B, Hijryana M, Akyea RK, Stokes J, Gibson J, et al. Eliciting and prioritising determinants of improved care in Multiple Long Term Health Conditions (MLTC): A modified online Delphi study. medRxiv [Internet]. 2023 Mar 19 [cited 2023 Mar 28];2023.03.19.23287406. Available from: https://www.medrxiv.org/content/10.1101/2023.03.19.23287406v1

27. Dambha-Miller H, Simpson G, Hobson L, Olaniyan D, Hodgson S, Roderick P, et al. Integrating primary care and social services for older adults with multimorbidity: a qualitative study. Br J Gen Pract [Internet]. 2021 Oct 1 [cited 2022 Dec 11];71(711):E753–61. Available from: https://pubmed.ncbi.nlm.nih.gov/34019480/

28. Carr E, Hagger-Johnson G, Head J, Shelton N, Stafford M, Stansfeld S, et al. Working conditions as predictors of retirement intentions and exit from paid employment: a 10-year follow-up of the English Longitudinal Study of Ageing. Eur J Ageing [Internet]. 2016 Mar 1 [cited 2023 Mar 12];13(1):39–48. Available from: https://link.springer.com/article/10.1007/s10433-015-0357-9

29. Torres JL, Lima-Costa MF, Marmot M, De Oliveira C. Wealth and Disability in Later Life: The English Longitudinal Study of Ageing (ELSA). PLoS One [Internet]. 2016 Nov 1 [cited 2023 Feb 4];11(11):e0166825. Available from: https://journals.plos.org/plosone/article?id=10.1371/journal.pone.0166825

30. Chudasama Y V, Khunti AK, Davies MJ. Clustering of comorbidities. Futur Heal J [Internet]. 2021 Jul 1 [cited 2022 Dec 11];8(2):e224–9. Available from: https://www.rcpjournals.org/content/futurehosp/8/2/e224

31. Fairley L, Cabieses B, Small N, Petherick ES, Lawlor DA, Pickett KE, et al. Using latent class analysis to develop a model of the relationship between socioeconomic position and ethnicity: Cross-sectional analyses from a multi-ethnic birth cohort study. BMC Public Health [Internet]. 2014 Aug 12 [cited 2022 Dec 11];14(1):1–14. Available from: https://bmcpublichealth.biomedcentral.com/articles/10.1186/1471-2458-14-835

32. van Blarikom E, Fudge N, Swinglehurst D. The emergence of multimorbidity as a matter of concern: a critical review. Biosocieties [Internet]. 2022 Sep 24 [cited 2023 Mar 28];1–18. Available from: https://link.springer.com/article/10.1057/s41292-022-00285-5

33. Robertson L, Vieira R, Butler J, Johnston M, Sawhney S, Black C. Identifying multimorbidity clusters in an unselected population of hospitalised patients. Sci Reports 2022 121 [Internet]. 2022 Mar 24 [cited 2023 Mar 28];12(1):1–10. Available from: https://www.nature.com/articles/s41598-022-08690-3

34. Pearson-Stuttard J, Ezzati M, Gregg EW. Multimorbidity—a defining challenge for health systems. Lancet Public Heal [Internet]. 2019 Dec 1 [cited 2023 Mar 28];4(12):e599–600. Available from: http://www.thelancet.com/article/S2468266719302221/fulltext

35. Henderson DAG, Atherton I, Mccowan C, Mercer SW, Bailey N. Linkage of national health and social care data: a cross-sectional study of multimorbidity and social care use in people aged over 65 years in Scotland. Age Ageing [Internet]. 2021 Jan 1 [cited 2023 Mar 28];50(1):176–82. Available from: https://pubmed.ncbi.nlm.nih.gov/32687158/

36. Busija L, Lim K, Szoeke C, Sanders KM, McCabe MP. Do replicable profiles of multimorbidity exist? Systematic review and synthesis. Eur J Epidemiol [Internet]. 2019 Nov 1 [cited 2022 Dec 11];34(11):1025–53. Available from: https://link.springer.com/article/10.1007/s10654-019-00568-5

37. Beach SR, Schulz R, Friedman EM, Rodakowski J, Martsolf RG, James AE. Adverse Consequences of Unmet Needs for Care in High-Need/High-Cost Older Adults. Journals Gerontol Ser B [Internet]. 2020 Jan 14 [cited 2022 Dec 11];75(2):459–70. Available from: https://academic.oup.com/psychsocgerontology/article/75/2/459/4869156

38. Prados-Torres A, Calderón-Larrañaga A, Hancco-Saavedra J, Poblador-Plou B, Van Den Akker M. Multimorbidity patterns: a systematic review. J Clin Epidemiol. 2014 Mar 1;67(3):254–66.

39. Ho HE, Yeh CJ, Wei JCC, Chu WM, Lee MC. Trends of Multimorbidity Patterns over 16 Years in Older Taiwanese People and Their Relationship to Mortality. Int J Environ Res Public Health [Internet]. 2022 Mar 1 [cited 2023 Mar 28];19(6). Available from: https://pubmed.ncbi.nlm.nih.gov/35329003/

40. Hernández B, Voll S, Lewis NA, McCrory C, White A, Stirland L, et al. Comparisons of disease cluster patterns, prevalence and health factors in the USA, Canada, England and Ireland. BMC Public Health [Internet]. 2021 Dec 1 [cited 2023 Mar 28];21(1):1–15. Available from: https://bmcpublichealth.biomedcentral.com/articles/10.1186/s12889-021-11706-8

41. Roman Lay AA, Ferreira Do Nascimento C, Caba Burgos F, Larraín Huerta ADC, Rivera Zeballos RE, Pantoja Silva V, et al. Gender Differences between Multimorbidity and All-Cause Mortality among Older Adults. Curr Gerontol Geriatr Res [Internet]. 2020 [cited 2023 Mar 28];2020. Available from: https://pubmed.ncbi.nlm.nih.gov/32148480/

42. Wang D, Li D, Mishra SR, Lim C, Dai X, Chen S, et al. Association between marital relationship and multimorbidity in middle-aged adults: a longitudinal study across the US, UK, Europe, and China. Maturitas. 2022 Jan 1;155:32–9.

43. Zhu Y, Edwards D, Mant J, Payne RA, Kiddle S. Characteristics, service use and mortality of clusters of multimorbid patients in England: A population-based study. BMC Med [Internet]. 2020 Apr 10 [cited 2022 Dec 11];18(1):1–11. Available from: https://bmcmedicine.biomedcentral.com/articles/10.1186/s12916-020-01543-8

