## Supplementary material file for "Clustering by Multiple Long-Term Conditions and Social Care Needs: A cohort study amongst 10,025 older adults in England"

**Supplementary Table 1:** Model fit and diagnostic criteria for competing latent class models.

|  | **LL** | **BIC(LL)** | **AIC(LL)** | **Npar** | **df** | **Classification Error** | **Dissimilarity index** | **Entropy R²** |
| --- | --- | --- | --- | --- | --- | --- | --- | --- |
| **1-Cluster** | -64952.8 | 130025.1 | 129931.7 | 13 | 8178 | 0 | 0.40 | 1 |
| **2-Cluster** | -61986.9 | 124221.9 | 124027.8 | 27 | 8164 | 0.0717 | 0.28 | 0.7401 |
| **3-Cluster** | -61733.4 | 123843.6 | 123548.8 | 41 | 8150 | 0.0736 | 0.27 | 0.7999 |
| **4-Cluster** | -61575.3 | 123656 | 123260.5 | 55 | 8136 | 0.1224 | 0.26 | 0.7194 |
| **5-Cluster** | -61421.7 | 123377.6 | 122781.5 | 69 | 8122 | 0.1688 | 0.25 | 0.6751 |
| **6-Cluster** | -61293.3 | 123449.3 | 122952.5 | 83 | 8108 | 0.2082 | 0.24 | 0.6605 |

**Abbreviations**: AIC, Akaike Information Criterion; BIC, Bayesian Information Criterion; df, degrees of freedom; LL, Log Likelihood; Npar, Number of parameters

**Supplementary Table 2:** Item response probabilities (95%CI) of the clusters of multiple long-term conditions and social care needs.

| **Cluster 1** | | **Cluster 2** | | **Cluster 3** | | **Cluster 4** | | **Cluster 5** | |
| --- | --- | --- | --- | --- | --- | --- | --- | --- | --- |
| **Condition/SCN** | **Probability** | **Condition/SCN** | **Probability** | **Condition/SCN** | **Probability** | **Condition/SCN** | **Probability** | **Condition/SCN** | **Probability** |
| Hypertension | 0.81 (0.75 - 0.86) | Difficulty with mobility | 0.85 (0.81 - 0.89) | Difficulty with mobility | 0.98 (0.94 - 0.99) | Difficulty with ADLs | 0.98 (0.94 - 0.99) | Difficulty with ADLs | 0.99 (0.92 - 1.00) |
| Mental health disorders | 0.37 (0.33 - 0.41) | Arthritis | 0.63 (0.59 - 0.66) | Arthritis | 0.51 (0.47 - 0.55) | Arthritis | 0.75 (0.72 - 0.77) | Difficulty with mobility | 0.98 (0.96 - 0.99) |
| Cardiovascular diseases | 0.34 (0.31 - 0.38) | Mental health disorders | 0.61 (0.58 - 0.64) | Health limit work | 0.49 (0.44 - 0.55) | Hypertension | 0.67 (0.65 - 0.69) | Health limit work | 0.80 (0.74 - 0.85) |
| Difficulty with mobility | 0.25 (0.14 - 0.42) | Health limit work | 0.46 (0.41 - 0.51) | Difficulty with ADLs | 0.38 (0.32 - 0.44) | Mental health disorders | 0.64 (0.62 - 0.66) | Hypertension | 0.65 (0.59 - 0.70) |
| Arthritis | 0.23 (0.20 - 0.27) | Cardiovascular diseases | 0.37 (0.34 - 0.40) | Mental health disorders | 0.36 (0.34 - 0.39) | Cardiovascular diseases | 0.50 (0.48 - 0.52) | Arthritis | 0.54 (0.48 - 0.60) |
| Lung condition | 0.23 (0.20 - 0.27) | Lung condition | 0.36 (0.33 - 0.39) | Cardiovascular diseases | 0.36 (0.33 - 0.38) | Lung condition | 0.36 (0.34 - 0.38) | Cardiovascular diseases | 0.53 (0.48 - 0.59) |
| Diabetes | 0.22 (0.19 - 0.25) | Difficulty with ADLs | 0.30 (0.25 - 0.35) | Diabetes | 0.22 (0.19 - 0.24) | Diabetes | 0.24 (0.22 - 0.26) | Dementia | 0.53 (0.38 - 0.67) |
| Health limits work | 0.16 (0.13 - 0.20) | Cancer | 0.28 (0.25 - 0.31) | Lung condition | 0.19 (0.16 - 0.21) | Cancer | 0.20 (0.19 - 0.22) | Mental health disorders | 0.46 (0.40 - 0.52) |
| Cancer | 0.15 (0.13 - 0.18) | Diabetes | 0.11 (0.09 - 0.14) | Cancer | 0.17 (0.16 - 0.19) | Stroke | 0.14 (0.12 - 0.16) | Stroke | 0.37 (0.32 - 0.43) |
| Stroke | 0.05 (0.04 - 0.07) | Stroke | 0.04 (0.03 - 0.05) | Stroke | 0.06 (0.04 - 0.07) | Dementia | 0.04 (0.02 - 0.06) | Diabetes | 0.23 (0.19 - 0.28) |
| Difficulty with ADLs | 0.02 (0.01 - 0.05) | Dementia | 0.02 (0.01 - 0.03) | Dementia | 0.00 (0.00 - 0.02) | Parkinson’s disease | 0.02 (0.01 - 0.03) | Cancer | 0.19 (0.15 - 0.24) |
| Parkinson’s disease | 0.00 (0.00 - 0.01) | Parkinson’s disease | 0.01 (0.00 - 0.02) | Parkinson’s disease | 0.00 (0.00 - 0.01) | Difficulty with mobility | 0.00 (0.00 - 0.00) | Lung condition | 0.19 (0.15 - 0.23) |
| Dementia | 0.00 (0.00 - 0.00) | Hypertension | 0.00 (0.00 - 0.00) | Hypertension | 0.00 (0.00 - 1.00) | Health limit work | 0.00 (0.00 - 0.00) | Parkinson disease | 0.10 (0.07 - 0.13) |

  *ADL is a composite of 13 variables = difficulty dressing, including putting on shoes and socks, difficulty walking across a room, difficulty bathing or showering, difficulty eating, such as cutting up food, difficulty getting in and out of bed, difficulty using the toilet, including getting up or down, difficulty using map to figure out how to get around strange place, difficulty preparing a hot meal, difficulty shopping for groceries, difficulty making telephone calls, difficulty taking medications, difficulty doing work around house and garden, difficulty managing money, e.g. paying bills, keeping track expense

**Mobility is a composite of 10 variables = difficulty walking 100 yards, difficulty sitting for 2 hours, difficulty getting up from a chair after sitting for long periods, difficulty climbing several flights of stairs without resting, difficulty climbing one flight of stairs without resting, difficulty stooping, kneeling or crouching, difficulty reaching or extending arms above shoulder level, difficulty pulling or pushing large objects, difficulty lifting or carrying weights over 10 pounds, difficulty picking up 5p coin from the table
